# Teamwork in the manufacturing workplace may hinder social distance

**DOI:** 10.1101/2020.08.22.20179994

**Authors:** Keisuke Kokubun

## Abstract

The spread of new coronavirus (COVID-19) infections show no signs of stopping. Therefore, if the era of With-Colona will continue for a while, we must consider how to deal with this disaster well. The practice of social distance is one of the powerful tools for that purpose. In previous research, analysis of the factors that influence social distance has also been carried out using information from the US occupation information site O*NET. However, they targeted all industries, not specific industries. Therefore, in this paper, we analyzed the factors that influence Physical Proximity in the manufacturing industry, which has a large impact on the economy given the scale of employment. As the method, first, exploratory factor analysis is performed using O*NET information, and the extracted 7 variables, Sitting Work, Adverse Conditions, Information Processing, Bridging Work, Teamwork, Response to Aggression, and Intelligent Work, are used in the regression analysis. As a result, it was shown that Teamwork is the biggest factor that influences Physical Proximity. Also, Adverse Conditions and Response to Aggression showed a positive correlation and Sitting Work showed a negative correlation. By job type, Maintenance, Installation & Repair tended to have higher Physical Proximity than Manufacturing Production Process Development, Production & Quality Assurance. Keywords: New Coronavirus (COVID-19), social distance, physical proximity, teamwork, explanatory factor analysis, O*NET

## Introduction

The threat of the new coronavirus (COVID-19) shows no signs of fading. As of August 20, 2020, the number of infected people in the world was 22,325,761 (Johns Hopkins University & Medicine, 2020). Until the development of the silver bullet, the “With-Colona” concept of how to coexist with this disaster is spreading. For coexistence, it will be necessary to continue to take the social distance that is currently recommended by the WHO in practice at the workplace (World Health Organization, 2020). As a result, researchers have been working over the past few months to analyze the factors that influence the feasibility of social distance and remote work (Crowley & Doran, 2020; Dingel & Neiman, 2020; Kokubun, 2020b; Koren & Pető, 2020). For example, a study by Kokubun (2020b) shows that it is the response to aggression that determines social distances most. Also, it is shown that there are the following occupations that require the response to aggression and physical proximity more: teachers of elementary/middle school and special education, therapists, technicians, nurses, employees of restaurants and entertainment facilities, travel/postal service clerks, flight/transportation attendants, etc. However, these studies are aimed at all industries, and as far as the author knows, there are no studies limited to specific industries. If the way people work differs depending on the industry, it is considered that the factors of social distance also differ between industries.

Therefore, this paper analyzes the factors that influence social distance, focusing on the manufacturing industry. As a method, first, following Kokubun (2020b), the factors common to 153 manufacturing-related occupations are extracted by exploratory factor analysis using the results of responses to 98 items recorded in Work Context and Work Activities of O*NET. Next, we aim to clarify the factors that influence social distance by performing regression analysis using variables created based on these factors as independent variables and Physical Proximity as dependent variables.

## Review of previous studies

Research on the prevention of coronavirus infection and its effect is increasing (e.g., Kokubun, 2020a; Kokubun et al., 2020). Relatedly, several studies have analyzed the factors that influence social distance (Crowley & Doran, 2020; Dingel & Neiman, 2020; Kokubun, 2020b; Koren & Pető, 2020). However, as far as the author knows, there is no analysis specialized for a particular industry. Given that people work differently in different industries, it will be necessary to analyze the factors that influence social distance in each industry. Among them, the manufacturing industry is considered to be of great significance for its research given its large impact on the national economy. For instance, in 2018, the number of workers employed in the manufacturing industry was 12,689 thousand, accounting for 7.9% of the number of workers in all industries in the United States (U.S. Bureau of Labor Statistics, 2019). We hope the analysis in this article will provide some helpful hints on what these workers need to do to maintain their social distance and work safety in the age of With-Colona.

## Method

Among questionnaire results posted on the O*NET website (https://www.onetonline.org/), we use the 98 items recorded in Work Context and Work Activities which were used in the studies of Koren & Pető (2020), Dingel & Neiman (2020), and others. Importance and Level are recorded in Work Activities, but Importance is used in the current research following previous research. All items are in the format of making a number from 0 to 100 for frequency and importance. In this paper, among 968 job types, we use answers from 153 job types that are classified as the manufacturing industry by Career Cluster (https://www.onetonline.org/find/career?c=13). Exploratory factor analysis is used to extract the factors that make up the variables. The criterion for factor extraction is eigenvalue 1 or more, and the factor load is calculated after performing varimax rotation by the main factor method. After that, items with a factor load of less than 0.4 and items with a factor load of 0.4 or more on a plurality of factors are excluded, and factor analysis is performed again using the same criteria. After that, this process is repeated until there are no items whose factor loads are less than 0.4 and items whose factor loads are 0.4 or more on plural items. Here, we follow the idea of Stevens (1992) who suggests using a cut-off of 0.4, irrespective of sample size, for interpretative purposes. After the factor structure is established, this paper presents a simple hypothesis that should be verified and performs a regression analysis using the variables consisting of each factor as the independent variable and Physical Proximity as the dependent variable to analyze the factors that influence social distance.

As a result of repeating the factor analysis 8 times by the above method, eight factors consisting of 44 items were extracted. However, The reliability coefficient of the factor consisting of two items of “Work Context-Importance of Being Exact or Accurate” and “Work Context-Time Pressure” was 0.544. This value is lower than the standard of 0.7 (Cortina, 1993) that many researchers show, or 0.6 (Taber, 2018; van Griethuijsen et al., 2015), which is a slightly looser standard. Therefore, it was determined that it would be difficult to use as a variable for analysis. Therefore, as a result of continuing the same processes after removing these items, 7 factors consisting of 40 items as shown in Table 1 were extracted in the 10th factor analysis. Note that the sentences listed are not the question text, but the content of the question text. Details of questions and options can be referred to on O*NET, so they are omitted in this article. However, to give an example, the question sentence corresponding to the content sentence at the top of the table “Work Context – Spend Time Standing” is “How much does this job require standing?”, and the options are “Never”, “Less than half the time”, “About half the time”, “More than half the time”, and “Continually or almost continually”. For each answer, 0 points, 25 points, 50 points, 75 points, and 100 points are assigned, and the average value is calculated for each occupation. Also, the Physical Proximity used as a dependent variable in the current research is selected from “I don’t work near other people (beyond 100ft.)”, “I work with others but not closely (e.g., private office)”, “Slightly close (e.g., shared office)”, “Moderately close (at arm’s length)”, and “Very close (near touching)” for the question sentence of “To what extent does this job require the worker to perform job tasks in close physical proximity to other people?” and is similarly assigned a value of 0 to 100 when totaling.

Based on the contents of the included items, the factors were named as Sitting Work, Adverse Conditions, Information Processing, Bridging Work, Teamwork, Response to Aggression, and Intelligent Work. Note that Teamwork is a little different from the Horizontal Teamwork of Kokubun (2020b). “Work With Work Group or Team” is an item that includes both horizontal and vertical relationships, while “Coordinate or Lead Others” and “Responsibility for Outcomes and Results” are items related to leadership. Since the manufacturing industry is an industry that requires various teamwork, such as between bosses and subordinates, or between colleagues, it can be interpreted that items with different contents are extracted as one factor. Similarly, Sitting Work, Bridging Work, and Intelligent Work are newly extracted factors in this analysis. On the other hand, the items that make up Adverse Conditions, Information Processing, and Response to Aggression largely overlap those of Kokubun (2020b). By the way, Physical Proximity is not included in any of these variables because the second factor analysis did not show loading of 0.4 or more for any factor.

**Table 1.** Results of exploratory factor analysis

| Item | Sitting<br>Work | Adverse<br>Conditions | Information<br>Processing | Bridging<br>Work | Teamwork | Response<br>to<br>Aggression | Intelligent<br>Work |
| --- | --- | --- | --- | --- | --- | --- | --- |
| Work Context - Spend Time Standing | **-0.902** | 0.147 | -0.028 | -0.050 | 0.091 | 0.156 | -0.008 |
| Work Context - Spend Time Sitting | **0.851** | -0.192 | 0.033 | 0.065 | -0.100 | -0.164 | 0.047 |
| Work Context - Spend Time Bending or Twisting the Body | **-0.842** | 0.231 | -0.109 | 0.052 | -0.098 | 0.081 | -0.058 |
| Work Context - Electronic Mail | **0.822** | 0.142 | 0.260 | 0.059 | 0.045 | 0.182 | 0.184 |
| Work Context - Telephone | **0.817** | 0.202 | 0.141 | 0.237 | 0.004 | 0.268 | 0.148 |
| Work Activities - Handling and Moving Objects | **-0.772** | 0.230 | -0.180 | 0.080 | -0.139 | -0.072 | -0.077 |
| Work Context - Letters and Memos | **0.717** | 0.241 | 0.124 | 0.071 | 0.239 | 0.174 | 0.066 |
| Work Context - Spend Time Walking and Running | **-0.695** | 0.272 | 0.049 | -0.187 | 0.083 | 0.312 | -0.098 |
| Work Context - Spend Time Using Your Hands to Handle, Control, or Feel Objects, Tools, or Controls | **-0.626** | -0.077 | -0.347 | 0.052 | -0.290 | -0.160 | -0.082 |
| Work Context - Spend Time Making Repetitive Motions | **-0.621** | -0.297 | -0.291 | 0.030 | -0.028 | -0.104 | -0.200 |
| Work Context - Outdoors, Exposed to Weather | 0.248 | **0.879** | 0.089 | 0.165 | 0.080 | 0.077 | -0.061 |
| Work Context - Exposed to High Places | 0.106 | **0.839** | 0.216 | -0.134 | 0.104 | 0.063 | -0.142 |
| Work Context - Outdoors, Under Cover | 0.313 | **0.832** | 0.113 | 0.176 | 0.063 | 0.047 | -0.043 |
| Work Context - Spend Time Climbing Ladders, Scaffolds, or Poles | -0.010 | **0.808** | 0.191 | -0.065 | 0.091 | 0.092 | 0.025 |
| Work Context - Cramped Work Space, Awkward Positions | -0.122 | **0.786** | 0.117 | -0.089 | -0.085 | 0.218 | 0.100 |
| Work Context - Extremely Bright or Inadequate Lighting | -0.132 | **0.768** | 0.156 | -0.126 | -0.021 | 0.104 | 0.001 |
| Work Context - In an Enclosed Vehicle or Equipment | 0.348 | **0.701** | 0.065 | 0.288 | -0.035 | 0.148 | 0.108 |
| Work Context - Very Hot or Cold Temperatures | -0.357 | **0.675** | 0.129 | -0.019 | 0.202 | 0.134 | -0.180 |
| Work Context - Exposed to Whole Body Vibration | -0.175 | **0.665** | 0.113 | 0.042 | 0.159 | -0.062 | 0.042 |
| Work Context - In an Open Vehicle or Equipment | -0.198 | **0.645** | 0.067 | -0.021 | 0.162 | -0.008 | -0.025 |
| Work Context - Wear Specialized Protective or Safety Equipment such as Breathing Apparatus, Safety Harness, Full Protection Suits, or Radiation Protection | -0.153 | **0.567** | 0.247 | -0.019 | 0.177 | -0.210 | -0.056 |
| Work Activities - Processing Information | 0.091 | 0.065 | **0.810** | 0.023 | 0.236 | 0.046 | 0.127 |
| Work Activities - Evaluating Information to Determine Compliance with Standards | 0.105 | 0.172 | **0.800** | -0.051 | 0.238 | 0.021 | 0.024 |
| Work Activities - Analyzing Data or Information | 0.367 | 0.093 | **0.779** | 0.102 | 0.120 | 0.016 | 0.234 |
| Work Activities - Monitor Processes, Materials, or Surroundings | -0.049 | 0.270 | **0.768** | -0.117 | 0.105 | -0.023 | -0.170 |
| Work Activities - Documenting/Recording Information | 0.309 | 0.226 | **0.735** | 0.018 | 0.166 | 0.108 | 0.004 |
| Work Activities - Identifying Objects, Actions, and Events | 0.026 | 0.291 | **0.733** | 0.037 | 0.010 | 0.001 | -0.065 |
| Work Activities - Making Decisions and Solving Problems | 0.272 | 0.184 | **0.696** | 0.261 | 0.011 | 0.089 | 0.328 |
| Work Activities - Getting Information | 0.172 | 0.060 | **0.642** | 0.007 | 0.134 | 0.078 | 0.208 |
| Work Activities - Selling or Influencing Others | 0.136 | -0.070 | 0.011 | **0.852** | -0.095 | 0.072 | 0.131 |
| Work Activities - Performing for or Working Directly with the Public | 0.101 | 0.043 | -0.139 | **0.840** | -0.246 | 0.079 | 0.026 |
| Work Activities - Staffing Organizational Units | -0.108 | 0.061 | 0.269 | **0.540** | 0.276 | 0.063 | -0.037 |
| Work Context - Work With Work Group or Team | 0.159 | 0.167 | 0.364 | -0.196 | **0.713** | 0.065 | -0.020 |
| Work Context - Coordinate or Lead Others | 0.283 | 0.217 | 0.284 | 0.003 | **0.711** | 0.163 | 0.059 |
| Work Context - Responsibility for Outcomes and Results | -0.090 | 0.217 | 0.259 | -0.039 | **0.667** | 0.193 | -0.018 |
| Work Context - Deal With Unpleasant or Angry People | 0.000 | 0.031 | 0.133 | 0.082 | 0.082 | **0.819** | -0.096 |
| Work Context - Frequency of Conflict Situations | 0.298 | 0.086 | 0.217 | -0.053 | 0.375 | **0.639** | 0.086 |
| Work Context - Deal With Physically Aggressive People | -0.070 | 0.136 | -0.059 | 0.106 | 0.062 | **0.616** | -0.050 |
| Work Activities - Drafting, Laying Out, and Specifying Technical Devices, Parts, and Equipment | 0.335 | -0.060 | 0.212 | -0.017 | 0.052 | -0.101 | **0.740** |
| Work Activities - Thinking Creatively | 0.287 | -0.139 | 0.187 | 0.356 | -0.051 | -0.098 | **0.579** |

Note(s): If the factor load is 0.4 or more, italic and bold type

## Hypothesis

Of the seven factors, Bridging Work, Teamwork, and Response to Aggression are factors that include relationships with people and are expected to require Physical Proximity. Indeed, it is stated, “physical proximity has a tremendous impact on the ability to work together” (Kiesler & Cummings, 2002, p.57). Besides, Adverse Conditions does not generally require closeness to people, but it is expected that some people will be required to have closeness if the job requires more than one person in case of an unexpected situation. From this, the following hypotheses are derived.

### H1

Adverse Conditions, Bridging Work, Teamwork, and Response to Aggression have a positive correlation with Physical Proximity.

On the other hand, Sitting Work, Information Processing, and Intelligent Work are considered to be jobs that do not require physical contact with people so often. From this, the following hypotheses are derived.

### H2

Sitting Work, Information Processing, and Intelligent Work have a negative correlation with Physical Proximity.

What has the strongest correlation with Physical Proximity? Sitting Work, Information Processing, and Intelligent Work are elements that do not require Physical Proximity strongly, and at the same time, they are not elements that actively distance Physical Proximity. Therefore, it is expected that any of the Adverse Conditions, Bridging Work, Teamwork, and Response to Aggression, which express relationships with people, is most strongly correlated with Physical Proximity. Of these, Adverse Conditions does not necessarily need to have multiple people if it is manual work. Bridging Work is also not difficult to implement remotely even in manufacturing enterprises if the information is enriched and clarified investing in information and communication technology (Swierczek & Kisperska-Moron, 2016; Townsend et al., 1998). Response to Aggression was the most important factor affecting Physical Proximity in the Kokubun (2020b) research targeting all industries. However, the manufacturing industry is less likely to deal with such people than the service industry, so the impact on Physical Proximity may not be so large. In this respect, Teamwork is an element required in many manufacturing industries. For instance, a previous study shows that team proximity in software development correlates with teamwork quality (Hoegl & Proserpio, 2004). Therefore, the following hypothesis is derived.

### H3

Teamwork has the strongest positive correlation with Physical Proximity.

## Analysis and result

Table 2 is descriptive statistics. Three variables, Adverse Conditions (r = 0.296), Teamwork (r = 0.393), and Response to Aggression (r = 0.309) showed a statistically significant positive correlation with Physical Proximity at the 1% level. However, the remaining Sitting Work, Information Processing, Bridging Work, and Intelligent Work did not show a significant correlation with Physical Proximity even at the 5% level. Table 3 shows the differences between industries. It is shown that there are differences among industries in 6 variables excluding Response to Aggression. The results of the Post-hoc Tukey HSD (Honestly Significant Difference) Test in the rightmost column show that there are significant differences in the values of these 6 variables between two or three industries. Therefore, in the following multiple regression analysis, we decided to use these industries after converting them into dichotomous values.

**Table 2.** Descriptive statistics

| | | Mean | SD | $\alpha$ | 1 | 2 | 3 | 4 | 5 | 6 | 7 |
| --- | --- | --- | --- | --- | --- | --- | --- | --- | --- | --- | --- |
| 1 | Physical Proximity | 57.390 | 10.293 |  |  |  |  |  |  |  |  |
| 2 | Sitting Work | 42.209 | 16.809 | 0.932 | -0.093 |  |  |  |  |  |  |
| 3 | Adverse Conditions | 25.141 | 14.422 | 0.928 | 0.296** | 0.051 |  |  |  |  |  |
| 4 | Information Processing | 63.996 | 9.635 | 0.928 | 0.120 | 0.376** | 0.375** |  |  |  |  |
| 5 | Bridging Work | 24.253 | 11.231 | 0.749 | -0.028 | 0.147 | 0.047 | 0.055 |  |  |  |
| 6 | Teamwork | 62.166 | 11.468 | 0.852 | 0.393** | 0.227** | 0.366** | 0.522** | -0.121 |  |  |
| 7 | Response to Aggression | 28.952 | 7.536 | 0.734 | 0.309** | 0.178* | 0.203* | 0.268** | 0.094 | 0.405** |  |
| 8 | Intelligent Work | 46.186 | 13.929 | 0.713 | -0.094 | 0.479** | -0.09 | 0.327** | 0.204* | 0.079 | -0.026 |

Note(s): n = 153; \*Significance at the 5% level; \*\*Significance at the 1% level.

**Table 3.** Differences between industries

|  | MPPD |  | MIR |  | PQA |  | F (2, 150) | Post-hoc Tukey HSD Test |
| --- | --- | --- | --- | --- | --- | --- | --- | --- |
|  | Mean | SD | Mean | SD | Mean | SD |  |  |
| Physical Proximity | 55.420 | 7.910 | 62.580 | 11.828 | 56.670 | 10.156 | 3.927* | 1-2*, 2-3* |
| Sitting Work | 64.108 | 8.104 | 49.621 | 9.598 | 34.954 | 13.996 | 60.679** | 1-2**, 1-3**, 2-3** |
| Adverse Conditions | 22.741 | 12.312 | 35.402 | 17.776 | 23.339 | 13.103 | 7.890** | 1-2**, 2-3** |
| Information Processing | 72.745 | 6.174 | 63.292 | 9.267 | 61.952 | 9.261 | 15.626** | 1-2**, 1-3** |
| Bridging Work | 21.436 | 8.049 | 31.792 | 13.355 | 23.207 | 10.741 | 7.218** | 1-2**, 2-3** |
| Teamwork | 67.885 | 6.332 | 58.944 | 17.193 | 61.473 | 10.404 | 4.573* | 1-2*, 1-3* |
| Response to Aggression | 30.859 | 7.424 | 30.847 | 7.160 | 28.029 | 7.545 | 2.408 |  |
| Intelligent Work | 60.385 | 13.285 | 48.958 | 8.241 | 41.956 | 12.622 | 24.531** | 1-2**, 1-3**, 2-3* |

Note(s): n = 153 (26 for MPPD; 24 for MIR; 103 for PQA);
MPPD = Manufacturing Production Process Development
MIR = Maintenance, Installation & Repair
PQA = Production & Quality Assurance
HSD = Honestly Significant Difference

Table 4 shows the results of regression analysis. Three industry variables and Seven independent variables are individually inputted into the regression equation. MIR (Maintenance, Installation & Repair), Adverse Conditions (β = 0.296), Teamwork (β = 0.393), Response to Aggression (β = 0.309) showed a significant positive correlation at 1% level, supporting H1. However, Sitting Work (β = –0.093), Information Processing (β = 0.120), and Intelligent Work (β = –0.094) did not show a statistically significant correlation even at the 5% level. Besides, Bridging Work (β = 0.120), which was expected to have a positive correlation with Physical Proximity, did not become significant at the 5% level. Comparing the adjusted R-squared, Teamwork was the largest at 0.149, followed by Response to Aggression at 0.090 and Adverse Conditions at 0.081. This shows that Teamwork has the greatest influence on social distance, and it can be said that this is the result of supporting H3. Industrial variables, MPPD (Manufacturing Production Process Development) and PQA (Production & Quality Assurance), did not become significant at the 5% level.

**Table 4.** Results of simple regression analysis with Physical Proximity as the dependent variable Variable

| Variable | $\beta$ | $R^2$ | Adj-$R^2$ | F |
| --- | --- | --- | --- | --- |
| MPPD | -0.087 | 0.007 | 0.001 | 1.140 |
| MIR | 0.219 ** | 0.048 | 0.041 | 7.572 ** |
| PQA | -0.100 | 0.010 | 0.003 | 1.529 |
| Sitting Work | -0.093 | 0.009 | 0.002 | 1.330 |
| Adverse Conditions | 0.296 ** | 0.087 | 0.081 | 14.356 ** |
| Information Processing | 0.120 | 0.014 | 0.008 | 2.205 |
| Bridging Work | -0.028 | 0.001 | -0.006 | 0.118 |
| Teamwork | 0.393 ** | 0.154 | 0.149 | 27.520 ** |
| Response to Aggression | 0.309 ** | 0.096 | 0.090 | 15.990 ** |
| Intelligent Work | -0.094 | 0.009 | 0.002 | 1.334 |

Note(s): n = 153; \*Significance at the 5% level; \*\*Significance at the 1% level.
MPPD = Manufacturing Production Process Development
MIR = Maintenance, Installation & Repair
PQA = Production & Quality Assurance

Table 5 shows the results of multiple regression analysis by the stepwise method. The first column shows the result of inputting only the dichotomous variables of three industries. MIR (β = 0.219, p<0.01) was selected as the statistically significant positively correlated variable, whereas MPPD and PQA were not selected. The second column is the result of inputting only 7 variables. Similarly to the results of simple regression, Adverse Conditions (β = 0.157, p<0.05), Teamwork (β = 0.302, p<0.01), Response to Aggression (β = 0.187, p<0.05) were statistically significant positive. This result supports H1. Also, as a result of controlling other variables, a significant negative correlation was shown in Sitting Work (β = –0.200, p<0.01). This result partially supports H2. However, Information Processing, Bridging Work, and Intelligent Work were not selected as significant variables in multiple regression as well as in single regression. The third column shows the results of an analysis conducted by adding three industrial-type variables to seven variables. Here, three variables of MIR (β = 0.330, p<0.01), Sitting Work (β = –0.265, p<0.01), Teamwork (β = 0.492, p<0.01) are significant positive correlations at 1% level. On the other hand, other variables including Adverse Conditions and Response to Aggression did not become significant at the 5% level. Comparing the coefficient of determination, 0.041, 0.217, 0.270 from the left, the model at the right end, the combination model of industrial-type variables and seven main variables, is the highest.

**Table 5.** Results of multiple regression analysis with Physical Proximity as the dependent variable Variable

| Variable | $\beta$ | | | |
| --- | --- | --- | --- | --- |
| MPPD |  | - |  |  |
| MIR | 0.219 ** | - | 0.330 ** |  |
| PQA |  | - |  |  |
| Sitting Work | - | -0.200 ** | -0.265 ** |  |
| Adverse Conditions | - | 0.157 * |  |  |
| Information Processing | - |  |  |  |
| Bridging Work | - |  |  |  |
| Teamwork | - | 0.302 ** | 0.492 ** |  |
| Response to Aggression | - | 0.187 * |  |  |
| Intelligent Work | - |  |  |  |
| R<sup>2</sup> | 0.048 | 0.237 | 0.284 |  |
| Adj-R<sup>2</sup> | 0.041 | 0.217 | 0.270 |  |
| F | 7.572 ** | 11.430 ** | 19.600 ** |  |

Note(s): n = 153; \*Significance at the 5% level; \*\*Significance at the 1% level. ‘-’ indicates that it is not used in the regression model. Blank cells indicate not selected in stepwise regression.
MPPD = Manufacturing Production Process Development
MIR = Maintenance, Installation & Repair
PQA = Production & Quality Assurance

For comparison, stepwise multiple regression analysis using 8 variables for all industries of Kokubun (2020b), i.e., Adverse Conditions, Leadership, Information Processing, Response to Aggression, Mechanical Movement, Autonomy, Communication with the Outside, and Horizontal Teamwork, were conducted. As a result, the statistically significant correlations were found in three variables, Response to Aggression (β = 0.218, p<0.01), Adverse Conditions (β = 0.186, p<0.05), and Horizontal Teamwork (β = 0.174, p<0.05). The adjusted R-squared was 0.147. The results of adding 3 industrial-type variables of to this are Response to Aggression (β = 0.193, p<0.05), Horizontal Teamwork (β = 0.295, p<0.01), Autonomy (β = 0.360, p<0.01), and MIR (β = –0.312, p<0.01) with the adjusted R-squared of 0.238. Cohen (1998) uses “effect size” criterion of 0.02 (small), 0.13 (medium), and 0.26 (large) for the scores of adjusted R-squared. Therefore, comparing the results in the third column, the adjusted R-squared shown by the model in this paper is large, and the adjusted R-squared when the all-industry model of Kokubun (2020b) is applied to the manufacturing industry is medium.

Figure 1 shows the correlation between Teamwork and Physical Proximity. The box in the lower left indicates that both Teamwork and Physical Proximity values are 0.5 standard deviations lower than the average value. The box in the upper right indicates that both Teamwork and Physical Proximity values are 0.5 standard deviations higher than the average value. Appendix A1 and A2 are extracted from the occupations in the lower left and upper right boxes for each variable. A1 (occupation with low Teamwork and low Physical Proximity) includes repairers of electronic/camera/medical/musical equipment, timing device assemblers/adjusters, computer-controlled machine tool operators, machine setters/operators/tenders of extruding/drawing/cutting/punching/press/welding/soldering/brazing, settlers/operators/tenders of textile bleaching/dyeing/knitting/weaving machine, etc. A2 (occupation with high Teamwork and high Physical Proximity) includes first-line supervisors of mechanics/installers/repairers and production/operating workers, meat cutters, food cooking machine operators/tenders, forging/rolling machine settlers/operators/tenders, model/pattern makers, system operators of a nuclear power reactor, power distribution, chemical plant, and petroleum pump, etc.

**Figure 1.**
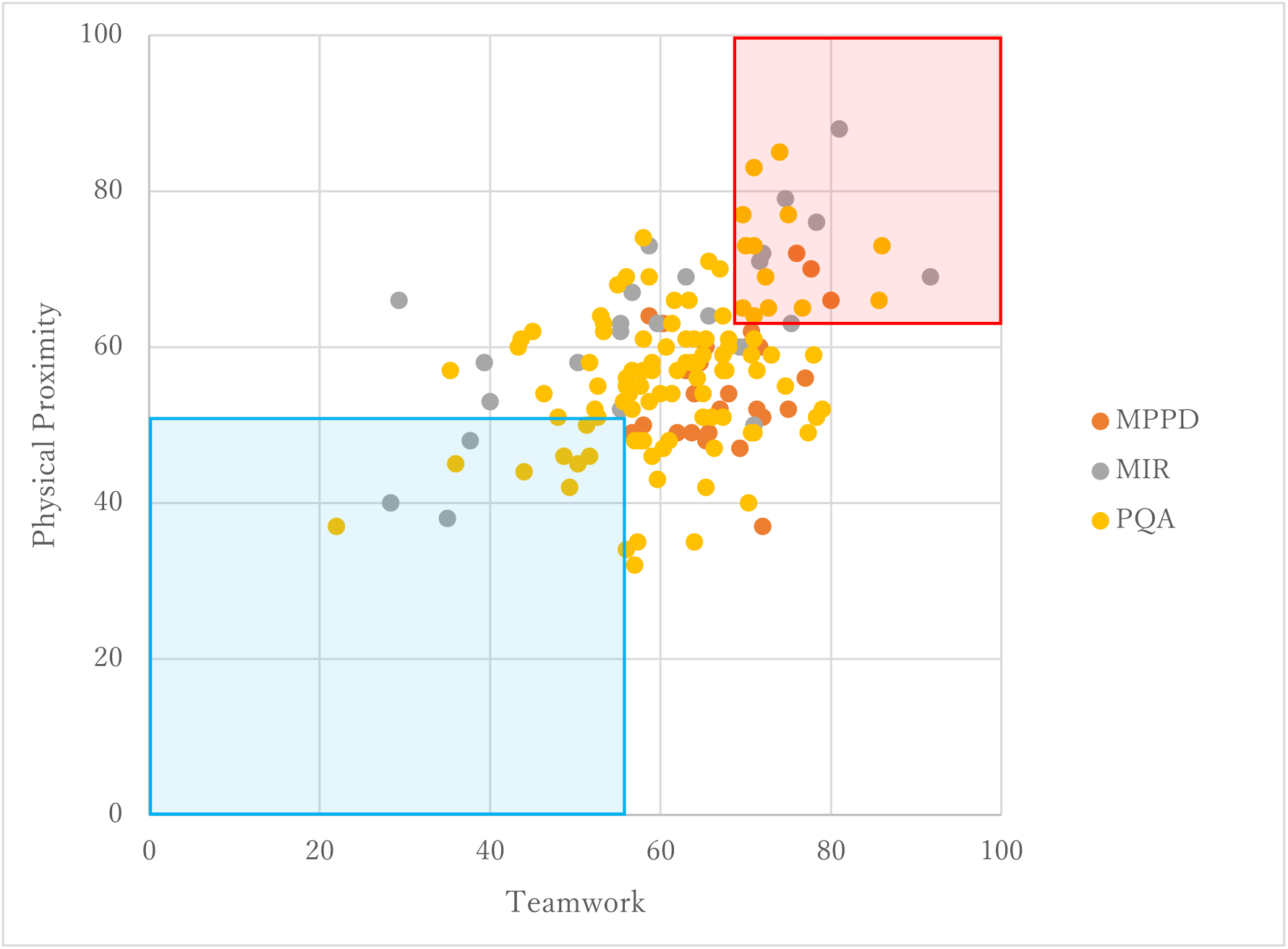
Scatter plot showing the relationship between Teamwork and Physical Proximity Note(s): The box in the lower left is 0.5 standard deviations lower than the average value. The box in the upper right is 0.5 standard deviations higher than the average value. MPPD = Manufacturing Production Process Development MIR = Maintenance, Installation & Repair PQA = Production & Quality Assurance

The former (A1) is characterized by the inclusion of repairers/assemblers/adjusters of some precision mechanical equipment and computer-controlled machine tool operators. It seems that teamwork and proximity are not required so much when working with craftsmanship, such as the repair or manufacturing of precision machinery, or when handling devices that are computer-controlled replacing human control. The latter (A2) is characterized by the inclusion of first-line supervisors and large-scale equipment system operators. It seems that teamwork and proximity are much required when dealing with people and huge devices. It is interesting to note that machine settlers/operators/tenders are included in both groups, even though they are of different types. This means that even when working with machines, there is a large difference in the degree to which teamwork and physical proximity are required.

## Discussion

The purpose of this paper was to find variables that correlate with Physical Proximity at the manufacturing site based on an exploratory method. The scales extracted as a result of the factor analysis were Sitting Work, Adverse Conditions, Information Processing, Bridging Work, Teamwork, Response to Aggression, and Intelligent Work. Of these, Adverse Conditions, Teamwork, and Response to Aggression were shown to have a positive correlation with Physical Proximity. This supports H1. Besides, as a result of multiple regression analysis, Adverse Conditions, Teamwork, and Response to Aggression showed a positive correlation and Sitting Work showed a negative correlation. Sitting Work did not correlate with Physical Proximity by itself, but a negative correlation was seen in multiple regression by controlling other variables such as Teamwork. It partially supports H2. Comparing the adjusted R-squared, it was also shown that Teamwork is the most important factor affecting Physical Proximity. This supports H3. On the other hand, Bridging Work, which was expected to have a positive correlation, and Information Processing and Intelligent Work, which were expected to have a negative correlation, did not show a significant correlation with Physical Proximity whether used for simple regression or multiple regression. This is inconsistent with H1 and H2.

Furthermore, even in a model in which 7 main variables and 3 industry variables were input at the same time, Teamwork showed a positive correlation and Sitting Work showed a negative correlation. In this case, the adjusted R-squared is 0.270, which is large according to the standard of Cohen (1988), and it showed improvement compared to the case of applying the model for all industries of Kokubun (2020b) to the manufacturing industry. By occupation, MIR (Maintenance, Installation & Repair) showed a statistically significant positive correlation with Physical Proximity, while PQA (Production & Quality Assurance) and HSD (Honestly Significant Difference) did not show a significant correlation. This result did not differ between simple regression and multiple regression. This means that the workers engaged in Maintenance, Installation & Repair have high Physical Proximity even if other differences such as Teamwork are controlled. In other words, it can be said that the practice of social distance is difficult for jobs that require Teamwork, jobs for which Sitting Work is difficult, and jobs for Maintenance, Installation & Repair, etc.

## Implication

In this paper, we analyzed the factors that influence Physical Proximity in the manufacturing industry by creating variables based on exploratory factor analysis and multivariate analysis using the information on questionnaire results recorded in O*NET, a job information website in the United States. As a result, it was shown that Teamwork showed the highest correlation with Physical Proximity. Also, the results showed that the job that is difficult to perform by Sitting Work and the job of Maintenance, Installation & Repair also require Physical Proximity. Teamwork has been found indispensable to the innovation (e.g., Montes et al., 2005) and employee commitment (e.g., Kokubun, 2018) of the manufacturing industry, and it is unlikely that this fact will change rapidly. However, in the With-Colona era, it is required to secure social distance as much as possible and continue production activities while preventing the spread of infection. For the occupations that require Teamwork and high Physical Proximity, as listed in Appendix Table A2, there is room to consider whether there is a method to secure social distance by changing the way of working while implementing infection protection measures. Besides, the required strength and contents of teamwork are considered to vary greatly depending on the workplace within the same industry. Efforts to devise ways to secure social distance, such as replacing existing face-to-face teamwork with virtual ones, are worth the effort. For example, some studies show that increasing employee trust affects virtual team performance more than face-to-face team performance (Breuer et al., 2016; Ford et al., 2017).

The variables extracted and created by the exploratory factor analysis for the manufacturing industry in this paper showed a higher correlation with the Physical Proximity than the variables extracted and created for all industries in Kokubun (2020b). This suggests that it is necessary to look at each industry to consider new ways of working towards securing social distance.

## Limitation

This paper exploratively extracted the factors that are the variables used in regression analysis, using the average values by the occupation of the attitude survey data recorded in the US occupation information site, O*NET. Therefore, if the primary data before being aggregated by occupation can be obtained and the results of this paper can be verified, its significance will be great. Besides, Physical

Proximity used as the dependent variable of the analysis is a variable based on the questionnaire survey results and may differ from the actual proximity. It is also significant to verify the analytical model in this paper after measuring the actual proximity using GPS location information, etc.

## Conclusion

The spread of new coronavirus (COVID-19) infections show no signs of stopping. Therefore, if the era of With-Colona will continue for a while, we must consider how to deal with this disaster well. The practice of social distance is one of the powerful tools for that purpose. In previous research, analysis of the factors that influence social distance has also been carried out using information from the US occupation information site O*NET. However, they targeted all industries, not specific industries. Therefore, in this paper, we analyzed the factors that influence Physical Proximity in the manufacturing industry, which has a large impact on the economy given the scale of employment. As the method, first, exploratory factor analysis is performed using O*NET information, and the extracted 7 variables, Sitting Work, Adverse Conditions, Information Processing, Bridging Work, Teamwork, Response to Aggression, and Intelligent Work, are used in the regression analysis. As a result, it was shown that Teamwork is the biggest factor that influences Physical Proximity. Also, Adverse Conditions and Response to Aggression showed a positive correlation and Sitting Work showed a negative correlation. By job type, Maintenance, Installation & Repair tended to have higher Physical Proximity than Manufacturing Production Process Development, Production & Quality Assurance.

## Data Availability

The raw data supporting the conclusions of this manuscript will be made available by the authors, without undue reservation, to any qualified researcher.

## Appendix

**A1.** Occupation with low Teamwork and low Physical Proximity

A1 Occupation with low Teamwork and low Physical Proximity
| Code | Occupation | Career Pathway | Teamwork | Physical Proximity |
| --- | --- | --- | --- | --- |
| 49-2097.00 | Electronic Home Entertainment Equipment Installers and Repairers | MIR | 28.3 | 40.0 |
| 49-9061.00 | Camera and Photographic Equipment Repairers | MIR | 35.0 | 38.0 |
| 49-9062.00 | Medical Equipment Repairers | MIR | 55.3 | 52.0 |
| 49-9063.00 | Musical Instrument Repairers and Tuners | MIR | 37.7 | 48.0 |
| 51-2093.00 | Timing Device Assemblers and Adjusters | PQA | 50.3 | 45.0 |
| 51-4011.00 | Computer-Controlled Machine Tool Operators, Metal and Plastic | PQA | 52.7 | 51.0 |
| 51-4021.00 | Extruding and Drawing Machine Setters, Operators, and Tenders, Metal and Plastic | PQA | 48.0 | 51.0 |
| 51-4031.00 | Cutting, Punching, and Press Machine Setters, Operators, and Tenders, Metal and Plastic | PQA | 51.3 | 50.0 |
| 51-4121.07 | Solderers and Brazers | PQA | 52.3 | 52.0 |
| 51-4122.00 | Welding, Soldering, and Brazing Machine Setters, Operators, and Tenders | PQA | 49.3 | 42.0 |
| 51-6051.00 | Sewers, Hand | PQA | 22.0 | 37.0 |
| 51-6061.00 | Textile Bleaching and Dyeing Machine Operators and Tenders | PQA | 48.7 | 46.0 |
| 51-6063.00 | Textile Knitting and Weaving Machine Setters, Operators, and Tenders | PQA | 44.0 | 44.0 |
| 51-9031.00 | Cutters and Trimmers, Hand | PQA | 56.0 | 34.0 |
| 51-9194.00 | Etchers and Engravers | PQA | 51.7 | 46.0 |
| 51-9195.05 | Potters, Manufacturing | PQA | 36.0 | 45.0 |

**A2.** Occupation with high Teamwork and high Physical Proximity

A2 Occupation with high Teamwork and high Physical Proximity
| Code | Occupation | Career Pathway | Teamwork | Physical Proximity |
| --- | --- | --- | --- | --- |
| 17-3029.03 | Electromechanical Engineering Technologists | MPPD | 80.0 | 66.0 |
| 19-4051.01 | Nuclear Equipment Operation Technicians | MPPD | 76.0 | 72.0 |
| 19-4051.02 | Nuclear Monitoring Technicians | MPPD | 77.7 | 70.0 |
| 49-1011.00 | First-Line Supervisors of Mechanics, Installers, and Repairers | MIR | 91.7 | 69.0 |
| 49-2021.00 | Radio, Cellular, and Tower Equipment Installers and Repairers | MIR | 78.3 | 76.0 |
| 49-2095.00 | Electrical and Electronics Repairers, Powerhouse, Substation, and Relay | MIR | 81.0 | 88.0 |
| 49-2098.00 | Security and Fire Alarm Systems Installers | MIR | 75.3 | 63.0 |
| 49-9041.00 | Industrial Machinery Mechanics | MIR | 72.0 | 72.0 |
| 49-9081.00 | Wind Turbine Service Technicians | MIR | 74.7 | 79.0 |
| 51-9082.00 | Medical Appliance Technicians | MIR | 71.7 | 71.0 |
| 51-1011.00 | First-Line Supervisors of Production and Operating Workers | PQA | 76.7 | 65.0 |
| 51-2091.00 | Fiberglass Laminators and Fabricators | PQA | 71.0 | 83.0 |
| 51-3021.00 | Butchers and Meat Cutters | PQA | 75.0 | 77.0 |
| 51-3022.00 | Meat, Poultry, and Fish Cutters and Trimmers | PQA | 74.0 | 85.0 |
| 51-3023.00 | Slaughterers and Meat Packers | PQA | 71.0 | 73.0 |
| 51-3093.00 | Food Cooking Machine Operators and Tenders | PQA | 70.0 | 73.0 |
| 51-4022.00 | Forging Machine Setters, Operators, and Tenders, Metal and Plastic | PQA | 72.7 | 65.0 |
| 51-4023.00 | Rolling Machine Setters, Operators, and Tenders, Metal and Plastic | PQA | 78.0 | 59.0 |
| 51-4061.00 | Model Makers, Metal and Plastic | PQA | 65.7 | 71.0 |
| 51-4062.00 | Patternmakers, Metal and Plastic | PQA | 67.3 | 64.0 |
| 51-8011.00 | Nuclear Power Reactor Operators | PQA | 86.0 | 73.0 |
| 51-8012.00 | Power Distributors and Dispatchers | PQA | 85.7 | 66.0 |
| 51-8091.00 | Chemical Plant and System Operators | PQA | 71.0 | 64.0 |
| 51-8093.00 | Petroleum Pump System Operators, Refinery Operators, and Gaugers | PQA | 72.3 | 69.0 |
| 51-8099.04 | Hydroelectric Plant Technicians | PQA | 69.7 | 77.0 |
| 51-9061.00 | Inspectors, Testers, Sorters, Samplers, and Weighers | PQA | 69.7 | 65.0 |
| Mean |  |  | 62.2 | 57.4 |
| SD |  |  | 11.5 | 10.3 |
| Mean + 0.5SD |  |  | 67.9 | 62.5 |
| Mean - 0.5SD |  |  | 56.4 | 52.2 |

